# Developing a Deep Learning Ultrasonography Model to Classify Thyroid Nodules as Benign

**DOI:** 10.1101/2022.12.31.22284087

**Authors:** Rishi Peddakama

**Affiliations:** Del Norte High School, San Diego, California, USA

## Abstract

1.

**<Introduction>:** Thyroid cancer, or the occurrence of rapid cell growth in the thyroid gland located near the neck, is the fastest growing cancer among women. Papillary thyroid cancer leads to hormonal imbalances thus causing periods of fatigue, difficulty breathing, and an overall decrease in one’s quality of life.

**<Objective>:** Unsurprisingly, the need for a quick diagnosis of thyroid cancer has become ever more important. Deep learning is a subset of machine learning that may improve the diagnostic performance of ultrasound, including in the evaluation of thyroid nodules (irregular growths that develop on the gland) for benign vs. malignant disease. Currently, the standard method to differentiate benign or malignant thyroid nodules is an invasive test known as Fine Needle Aspirations (FNAs) that is uncomfortable and may lead to unwanted outcomes such as bleeding, and infection.

**<Methods>:** We propose using a convolutional neural network with a foundation rooted in two pre-trained networks, VGG-16 and InceptionV3, to train a model on the publicly available Thyroid Digital Image Database to accurately classify thyroid nodules on ultrasonography into those that are malignant or benign; a non-invasive strategy.

**<Results>:** The data was augmented using industry-standard methods and provisioned into train/test/validation sets obtaining 88% accuracy and an area under the receiver operating characteristic (ROC) of 0.929 (signifying a high sensitivity).

**<Discussion>:** Based on the model’s results, it is conceived that a convolutional neural network can serve as an accurate classifier of malignant/benign thyroid nodules from an ultrasound and enhance a diagnostic strategy to reduce unnecessary FNA procedures.

## 2. INTRODUCTION

The thyroid is a butterfly-shaped organ located in the human neck responsible for the secretion of two vital metabolic hormones: triiodothyronine (T3) and thyroxine (T4). Thyroid nodules are solid or fluid-filled lumps within the thyroid that are classified as either benign (noncancerous growth) or malign (cancerous growth) [10]. Though they may sometimes grow to a visible size or make it difficult to swallow and breathe, thyroid nodules are often only detected during medical exams or scans. These nodules, if allowed to grow rapidly, can become cancerous and thereby hinder the function of a vital organ to the human endocrine system. A loss of thyroid functionality and the subsequent hormones it produces causes hypothyroidism characterized by sluggishness, decreased metabolism, and other health risks [11]. Additionally, if failed to be treated properly, thyroid cancer can spread to other parts of the body primarily (and dangerously) the lungs. Thyroid cancer affects over 45,000 people in the United States annually and is the fastest growing cancer among women [18]. This, coupled with its difficulty to detect in early stages proves that research must be done to improve the diagnosis process.

Diagnosis first begins with a physical examination from an endocrinologist in which patients are looked at for abnormally sized lumps near the neck and asked to swallow to see how it changes. Then, physicians may use a plethora of diagnostic techniques for identifying thyroid nodules namely Fine Needle Aspirations (FNA) which involves sampling tissue from the thyroid gland and sending them to the lab for analysis. The results provide information on the quantity of triiodothyronine and thyroxine is being produced by the thyroid [7]. Abnormal amounts of either hormone signify an abnormality and the possible existence of malignant nodules. Despite being an effective method in thyroid cancer diagnosis, FNAs are invasive, painful (that is, you have to get stuck with a needle in the lower neck area), and time-consuming. Not only that, but a patient must take precautions many days before the procedure by watching their diet, and the consumption of certain medications such as blood thinners (which might be important for one’s cardiac health). There also exists image-based ultrasound analysis for diagnosis. Ultrasound imaging, also known as sonography, is the practice of sending sound waves through a certain region of the human body [1]. These sound waves, when coming into contact with boundary-like objects (such as tissue, liquid, or bone) bounce off them and return to the probe [1]. With the aid of a computer system, the information on each wave’s speed, time, and direction can be analyzed and represented as a 2-D grayscale image. The higher the intensity of a wave, the lighter it appears and vice versa. Unlike FNAs, ultrasounds provide physicians with visuals of the size of the tumor and its location. Thus, ultrasounds are very beneficial tools in the realm of tumor identification and cancer diagnosis. A significant drawback of ultrasounds, however, is that they require a medical expert to identify benign/malign nodules thereby making it a time-consuming and resource-intensive task (one must contact a radiologist specializing in thyroid cancer and wait for their diagnosis). The thyroid’s importance to the human body is the reason why easy treatment and diagnosis of benign and malignant nodules is becoming increasingly more important for thyroid cancer. Wouldn’t it be great if thyroid ultrasounds could be analyzed to evaluate nodules leading to less pain and less risk of infection? Enter Computer-Aided Diagnosis (CAD) and its ability to make decisions swiftly and cheaply [5].

The advent of digital technology in the medical field has paved the way for doctors to analyze diseases not visible to the naked eye. Of these technologies, ultrasounds have been analyzed in various applications of medicine as a way of gaining a holistic understanding of various diseases. However, these ultrasounds often require an expert radiologist to visually analyze them and evaluate them for cancers. Fortunately, the advancements in GPU technologies have made these images very useful when combined with Convolutional Neural Networks (CNNs) and other deep learning architectures. Ultrasounds can now be analyzed in mere seconds with deep learning models no longer needing the time and possible human error of medical experts. A deep learning application of thyroid ultrasounds provides a novel approach to make diagnosis more accurate and consistent. We employed a model that analyzes these images synthesized from high-frequency sound waves to accurately and quickly identify thyroid nodules. The objective of this pilot study is to determine if a CNN can be trained to identify if a thyroid ultrasound contains benign or malignant nodules.

## 3. METHODS

### 3.1 Dataset

The Thyroid Digital Image Database (TDID), a public dataset published in 2015 by the Universidad Nacional de Colombia contains ultrasound thyroid images of patients [14]. Each image, with an average dimension of 560 × 315 pixels, was sorted into categories by radiologists using the Thyroid Imaging Reporting And Data System (TI-RADS). The five suspicion levels were: benign (TR1), not suspicious (TR2), mildly suspicious (TR3), moderately suspicious (TR4), and highly suspicious (TR5). Of the groups given by the dataset (TR2, TR3, TR4, TR5), TR2 and TR3 ultrasounds were classified as benign while TR4 and TR5 were labeled malignant [15, 16].

#### 3.1.1 Thyroid Imaging Reporting and Data System

With nearly 67% of the US population having thyroid nodules [10] (most of which are benign), radiologists felt the need to establish a set of guidelines for accurate and consistent treatment of thyroid-related cancers [16]. Subsequently, the American College of Radiology designed the Thyroid Imaging Reporting and Data System (TI-RADS) to improve overall diagnostic accuracy. Thyroid nodules could now be sorted into 5 categories based on analytics received from an ultrasound: composition, echogenicity, shape, margin, and punctate echogenic foci. Certain combinations of TR-classifications along with length determine if malignant nodules require clinical attention or if Fine Needle Aspirations (FNAs) are applicable [15, 16].

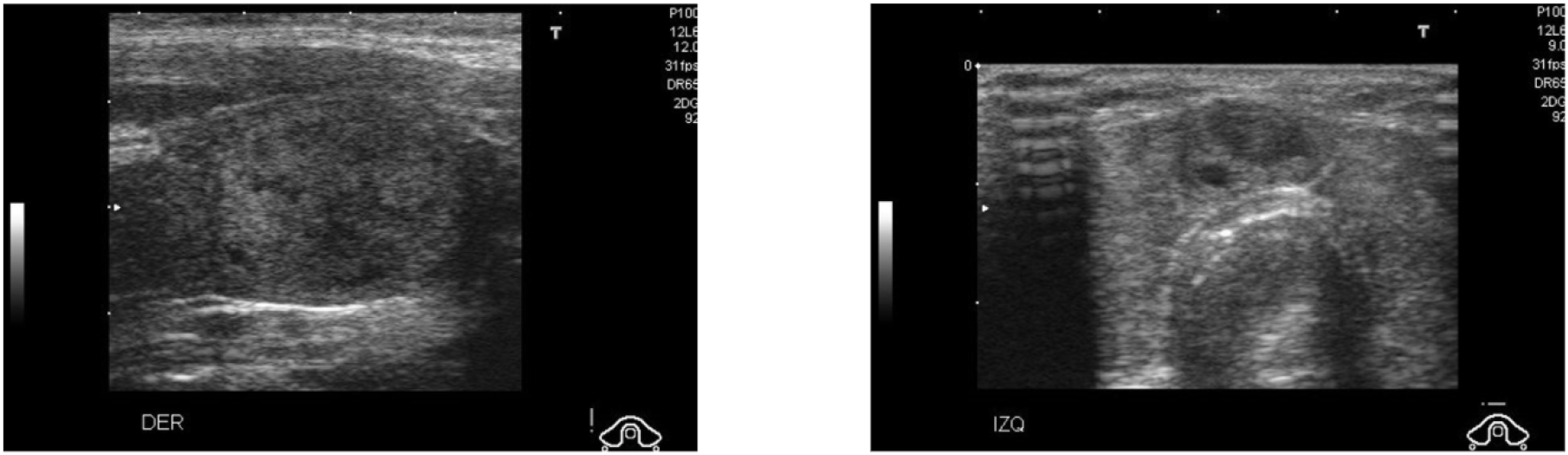

As mentioned by ultrasound characteristics before, lighter regions represent more pronounced characteristics of the region while darker regions are part of the background and are closely related to “image noise”. Purely from visual observations, it is evident that malignant nodules (TI-RADS 4 image on the right) have more defined circular areas compared to the benign nodules (TI-RADS 2 image on the left). It is this differentiation that we hope to replicate with a deep learning model.

### 3.2 Preprocessing and Augmentation

First, the images were normalized by file size in order to fit a neural network structure. The data was split in the following way:

- Training Images (used to tune CNN hyperparameters): 55 Malignant, 13 Benign (68 total).
- Testing Images (used to tune CNN hyperparameters): 7 Malignant, 1 Benign (8 total).
- Validation Set (serves as a “hold out” and does not influence training): Validation split set to 0.1 of the entire dataset

The dataset was relatively small and thus to enhance training, image augmentation was employed to create more training samples. Early stopping was also added before compiling the model to halt overfitting. Prior to being fed into the network, the pixel values for all images were normalized to values between 0 and 1 by simply dividing by 255 (the max grayscale value). Without doing this, gradient descent (or the actual training part) will be greatly affected by weights that are too large to account for large pixel values. Additionally, most activation functions such as tanh and sigmoid are designed to output values between −1 and 1, and inputting large numbers like 255 into these functions will produce values very close to 1 each time effectively making that particular node’s weight worthless.

#### 3.2.1 Image Augmentation

Because its learning capabilities are rooted in gradient descent, deep learning is known to become increasingly more accurate with a greater amount of data. However, the TDID dataset, with few images it provides, certainly hinders the model’s potential. Image augmentation is the process of altering existing datasets to artificially diversify a dataset. Many of the common techniques include image rotation, image shifting, image flipping, image noising, and image blurring [3]. For this research’s application, we chose to use all methods except image noising and image blurring. Ultrasounds, by default, already have noise represented by darker shaded areas and spotty picture quality (known as Speckle noise; it decreases the overall resolution of the image [3]). Blurring a black and white image would push a model to become a randomized classifier as the ultrasounds would become too distorted beyond any noticeable regions.

### 3.3 Addressing Overfitting with Early Stopping

In deep learning, when loss functions produce values beyond a threshold set by the researcher, training is stopped at that instant to save computing power and prevent overfitting [12]. In gradient descent terms, as the slope of the axis begins to flatten out greatly, there is no longer a need for backpropagation. Early stopping is determined by validation loss (as this is not part of the training process). Choosing the threshold for a particular model depends on a variety of factors including the size of the dataset, batch size, and epochs [12]. The main determiner of this value is the computing power of the system the model is being trained on. The more processing power, the lower the threshold can be and vice versa.

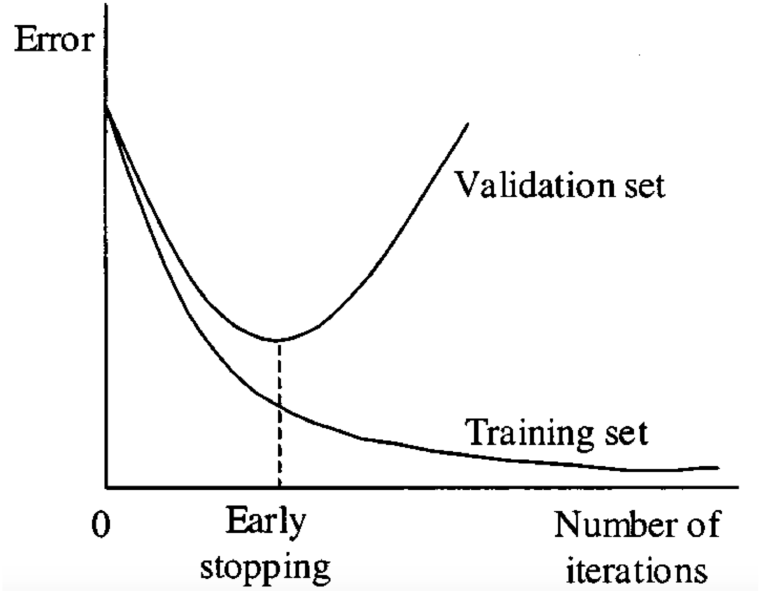

In the image above [12], as error begins to plateau in the quadratic function, training likely ends at this point as each epoch brings little change to the accuracy of the model.

### 3.4 Data Distribution

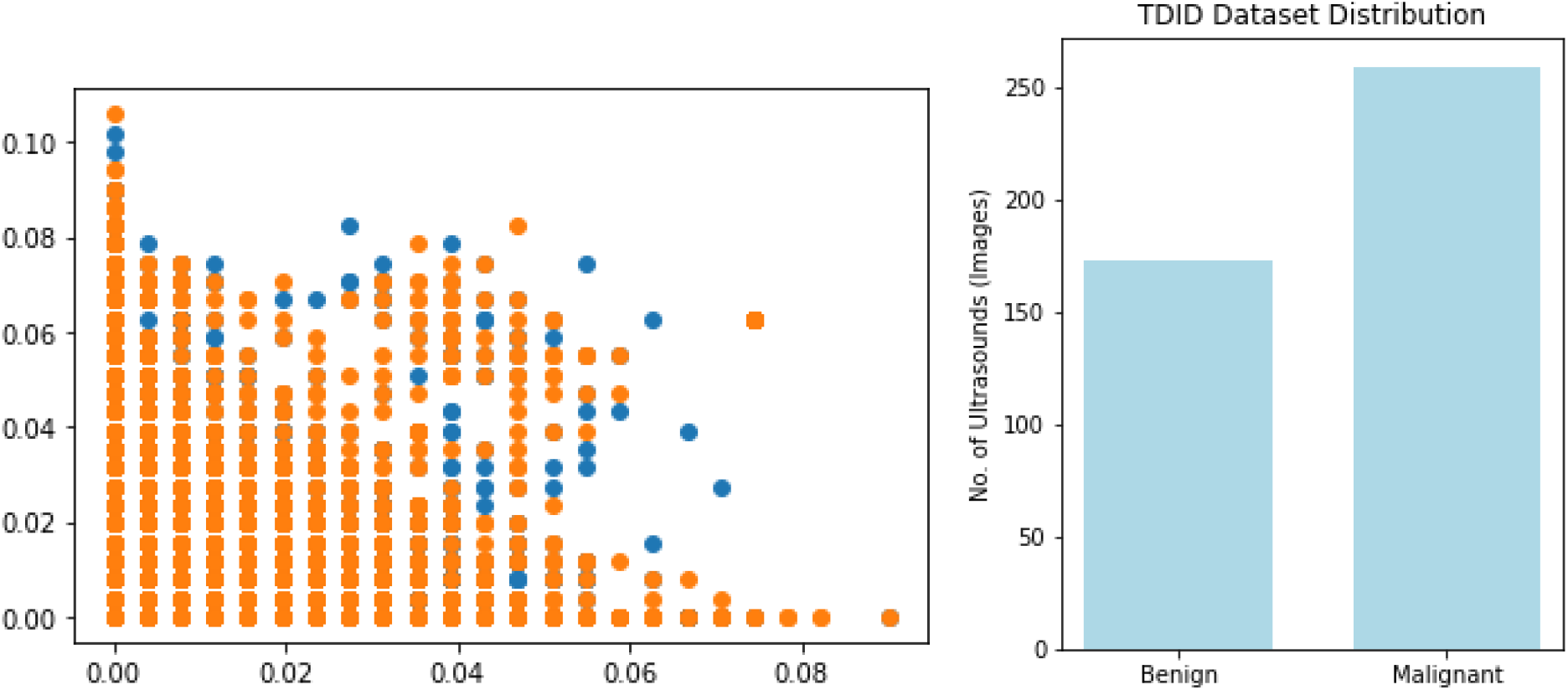

The figures above represent the distribution of classes in the TDID dataset. Evidently, Malignant ultrasounds appeared far more frequently. To avoid overfitting, class weights were initialized before training the CNN to favor Benign cases by a factor of roughly 4 (Further addressed later).

### 3.5 Robust CNN Model Creation

#### 3.5.1 Deep Learning

Deep learning is the use of a multi-layered neural network for the classification of various kinds of data including photo, video, and audio data. A neural net’s architecture was first conceptualized back in 1944 by Warren McCullough and Walter Pitts from the University of Chicago marking one of the earliest crossings of the field of neuroscience and computer science [4]. To put it simply a neural network is modeled on the human brain with thousands of nodes interconnected with each other layer by layer. In the most common model pass, feed-forward, data moves in a single direction and is multiplied by the individual weights of the nodes in a matrix matter. In the final layer (the number of nodes is determined by the number of outputs needed for the specific classification task), the highest number is deemed the “answer” and compared with the training case [4]. Generally, all node values in the last layer are between 0 and 1 to prevent skewed calculations. Then, by calculating loss based on the correct train output (there are a variety of loss functions that each have their purpose), backpropagation is used to fine-tune weights. Over time, with a large amount of data, the neural net’s nodes become fine-tuned enough to accurately classify images [4]. Despite being discovered nearly 80 years ago, neural networks have had little development until recently with the development of fast, affordable GPUs that make training times exponentially shorter.

#### 3.5.2 Convolutional Neural Network

Recently, a category of neural nets, Convolutional Neural Networks (CNNs), has become very popular recently because of its application to computer vision and image classification. CNNs’ popularity skyrocketed following the contributions to the ImageNet Large Scale Visual Recognition Competition (ILSVRC) in 2012 [9, 19]. CNNs are characterized by special layers: convolution, pooling, and fully connected layers [19]. To train the model, images are transformed into 2D arrays of their pixel values; numbers between 0-255 (though they are typically normalized to a value between zero and one to prevent certain weights from becoming too biased). If the images are RGB, then each image will consist of three 2D arrays representing red, green, and blue hues respectively. In each Convolution, the image is iterated over by a grid known as a kernel. Because kernels can have varying dimensions, this makes convolutional neural networks very fast as high-resolution images no longer have to be iterated over pixel by pixel but can be viewed as multiple “snapshots”. After each layer has its distortions removed with max-pooling and is passed through an activation function (most notably Rectified Linear Unit or ReLU) and the output is received, the machine trains itself with backpropagation and gradient descent.

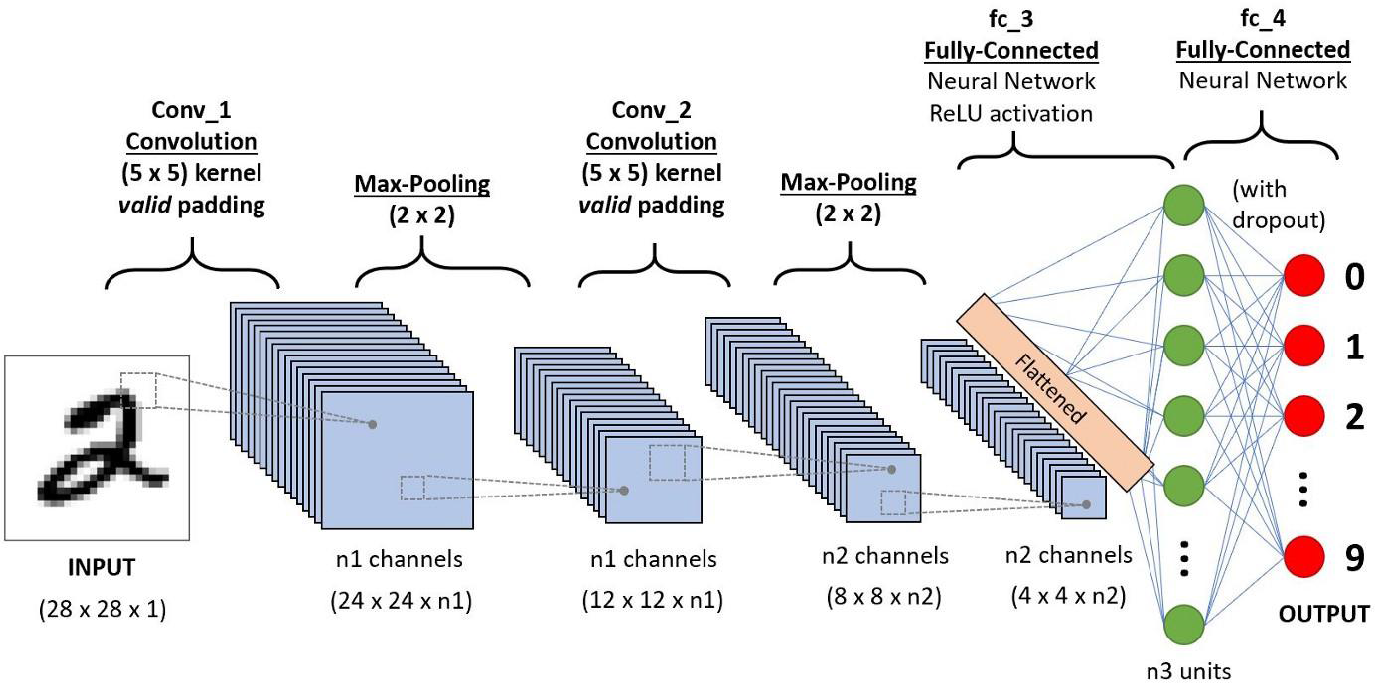

The image above represents the general architecture of a convolutional neural network, the best deep learning model for image classification.

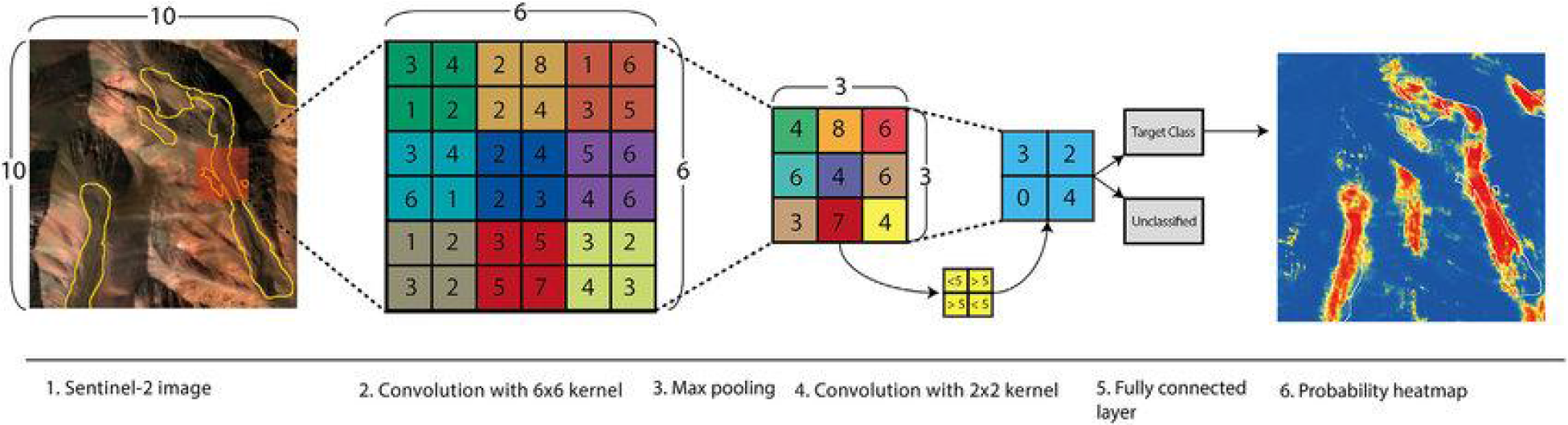

The figure above [13] represents the step-by-step process a CNN takes during an iteration.

#### 3.5.3 Tools

A desktop computer was used to train the deep learning model. It had an Intel Core i7-9700 CPU and an NVIDIA GeForce GTX 1660 Ti graphics card. 16 gigabytes of RAM proved sufficient for the size of the proposed model. The convolutional neural network itself was implemented using Python and the Tensorflow library. As mentioned, the model’s architecture requires the weights of 2 pre-trained models, both of which are also from the Tensorflow library. Other libraries were used for data preprocessing, visualization, and other statistics. These were: Numpy, Pandas, Pillow, Matplotlib, Sklearn, Seaborn.

#### 3.5.4 Architecture

Traditionally, deep learning requires large amounts of data to properly tune the parameters of a given neural network. The acquisition of medical data, specifically ultrasounds, is significantly harder due to the legal constraints and privacy policies of medical institutions. Alternatively, pre-trained CNNs, trained on enormous amounts of data and designed for general image classification, serve as a perfect backbone for computer vision models. A combination of two popularized image classification networks (VGG16, InceptionV3) was proposed for this diagnostic task.

#### 3.5.5 Very Deep Convolutional Networks for Large-Scale Image Recognition (VGG-16)

VGG16 is a convolutional neural network model invented by A. Zisserman and K. Simonyan from the University of Oxford. When tested on ImageNet, a dataset of over 14 million images from 1000 distinct classes, the model performed with 92.7% accuracy. The model takes in an input shape of 244×244×3 (though this is later changed to fit the images in the TDID dataset) and has a series of carefully picked out and sized convolution, max-pooling, and fully-connected layers. Because of its success in such a wide range of image categories, VGG-16 is often used as a base in many deep learning projects as the CNN’s weights are already fine-tuned for general image analysis. The image below is the structure of VGG-16 [17]

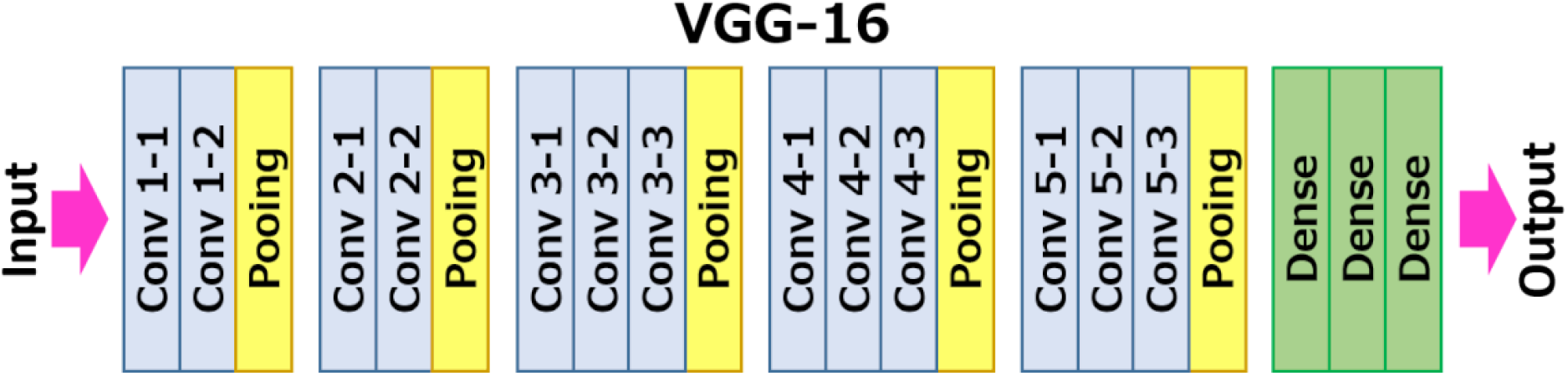

### 3.5.6 Inception v3

Inception v3 is also a popular convolutional neural network. It has 78.1% accuracy on the ImageNet dataset but combines many new research ideas for neural nets including symmetric and asymmetric building blocks [6]. We chose to add Inception v3 in addition to VGG-16 because of past applications of Inception v3 in medical imaging. The combination of these two models, from the high accuracy for general images of VGG-16, to the specially-tailored for medicine Inception v3, will create a strong classifier of thyroid nodules from ultrasound images. An image of Inception v3’s architecture is below [6].

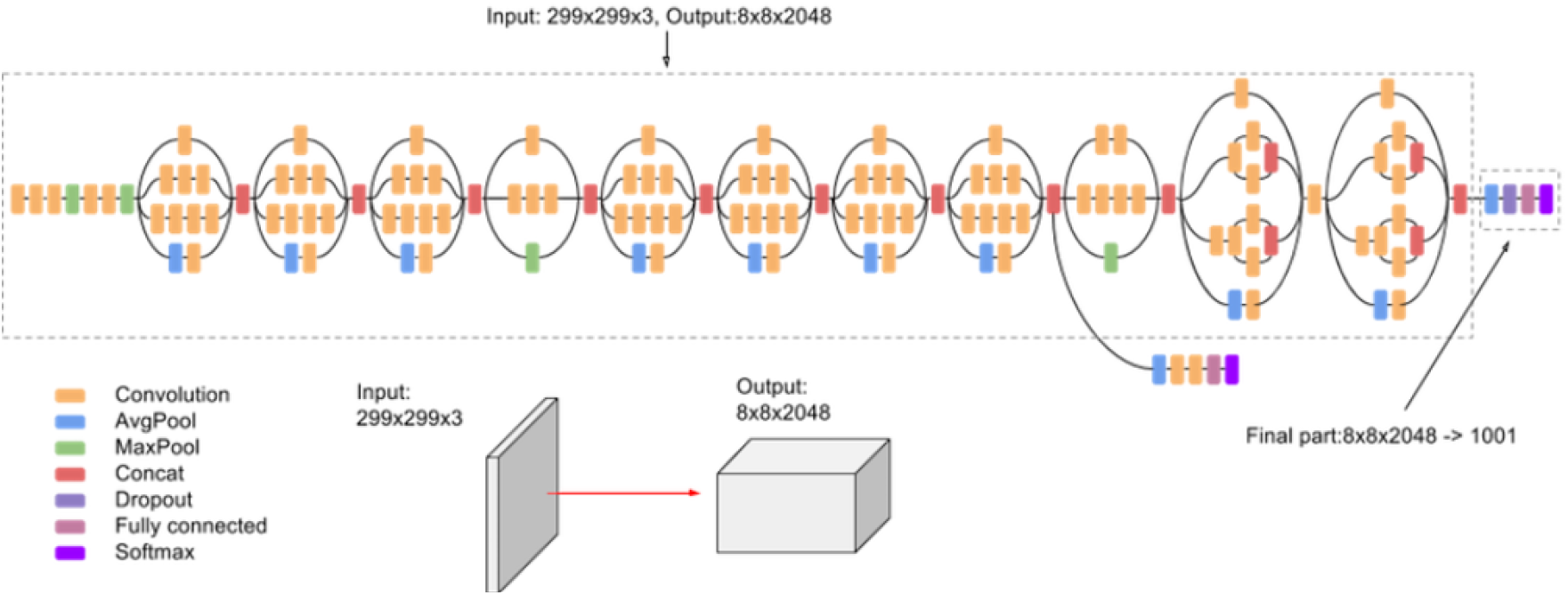

Because the VGG16 model has been tailored to identify images from a vast number of categories, it lacks the specificity to be used as the only model for ultrasound imaging. InceptionV3 on the other hand, with its unique structure of a parallel max-pooling layer, adds depth to the model. The figures below contain the holistic architectures of both these models [17].

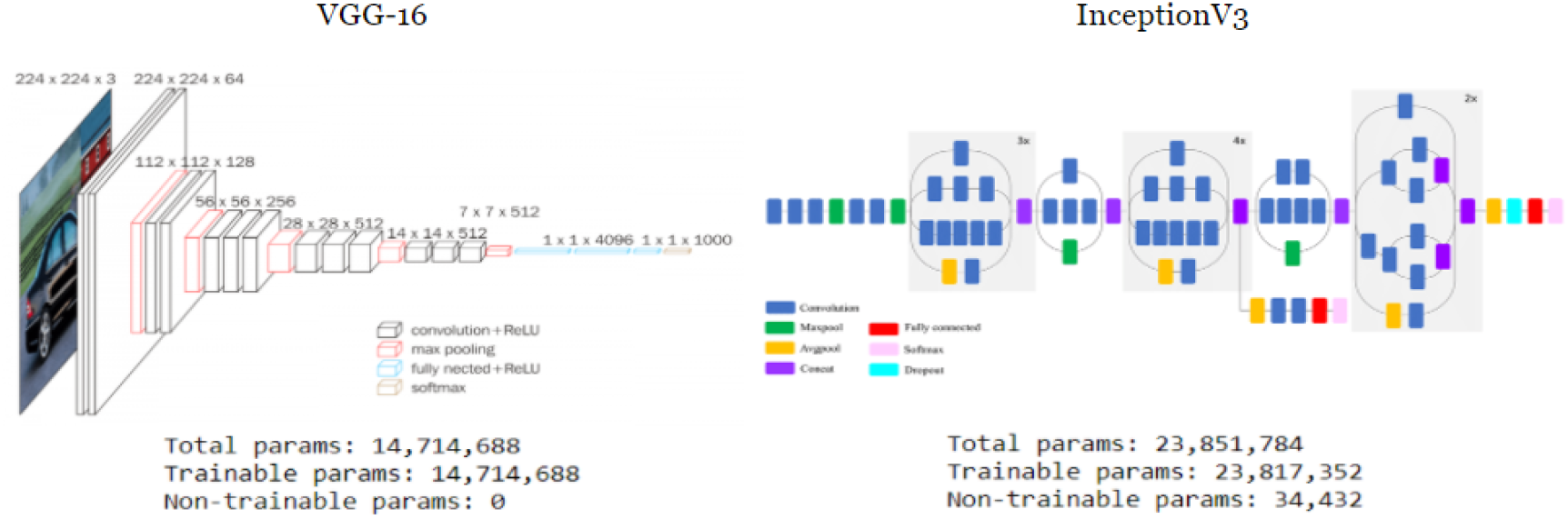

In the model diagram above, a thyroid nodule ultrasound is received as input with the neural network outputting a single node (a positive or negative test for malignancy). Because VGG16 is designed to take in RGB with an input shape of <u>width x height x 3</u>, the ultrasound images were tripled to match this structure.

#### 3.5.7 Loss Function

In most classification models, there are a select amount of loss functions to choose from. The choice is narrowed down based on factors such as output size, image type, and data skew. For this research, since the model is a binary classification of either malignant or benign nodules, binary cross-entropy is the clear answer. However, this isn’t enough; because the TDID has a far greater amount of malignant ultrasounds (roughly 4x more), the model was fitted with weights favoring benign images 4 times more than malignant ones.

#### 3.5.8 Addressing Unbalanced Data

In many public datasets, it is common to encounter an unbalanced amount of samples among different classes. In order to counter this, class weights can be preset to value the prediction error of the smaller class far more than that of a larger class. A simple ratio was constructed to find the proportion of benign images in the entire dataset which was made out to be approximately 4. Thus, while training, errors that related to classifying benign images were modified by a factor of 4. Doing this effectively removed the problem of unbalanced data making a more cohesive model.

#### 3.5.9 Binary Cross-Entropy

Binary cross-entropy, as evident by its name, provides an output with only 2 possible choices.

The formula for the function is as follows: (Where yhat is the model’s output and y is the target output)

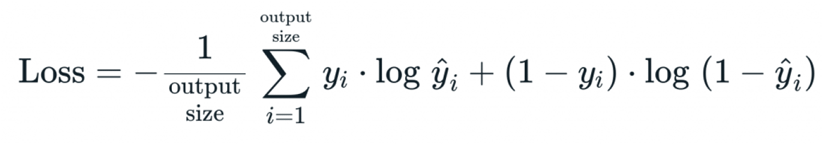

The sigmoid activation function works best with binary cross-entropy and is applied at the end of the output layer [2]. The function outputs a value between 0 and 1. When rounded, this value can only represent two values hence why it is so useful for binary cross-entropy [2].

## 4. RESULTS

### 4.1 Metrics

Generally, deep learning models are evaluated on many metrics, the notable ones being specificity, sensitivity, and accuracy. These metrics are especially important for this research as the model focuses on thyroid cancer diagnosis.

#### 4.1.1 Sensitivity

Sensitivity is the ratio between true positive cases (in this experiment, that being ultrasounds of malignant thyroid nodules that the model correctly predicted as being malignant) and the number of positive data values. The formula can be represented as (#True Positives)/(#True Positives + #False Negatives) values. It is also known as the true positive rate (TPR). For most deep learning models, especially those that have an application to the medical field, high sensitivity values are desirable as they signify high accuracy when it comes to diagnosis and reduces the need for unnecessary procedures on false positives.

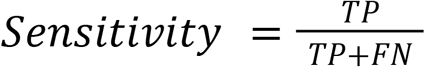

#### 4.1.2 Specificity

Specificity is the ratio between true negatives (in this experiment, that being ultrasounds of benign thyroid nodules that the model correctly predicted as being benign) and the number of negative data values. This is mathematically represented as (#True Negatives)/(#True Negatives + #False Positives). Specify is also referred to as the true negative rate (TNR). In medicine, high specificity is also very important to prevent cases where a disease goes undiagnosed in a patient.

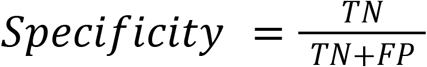

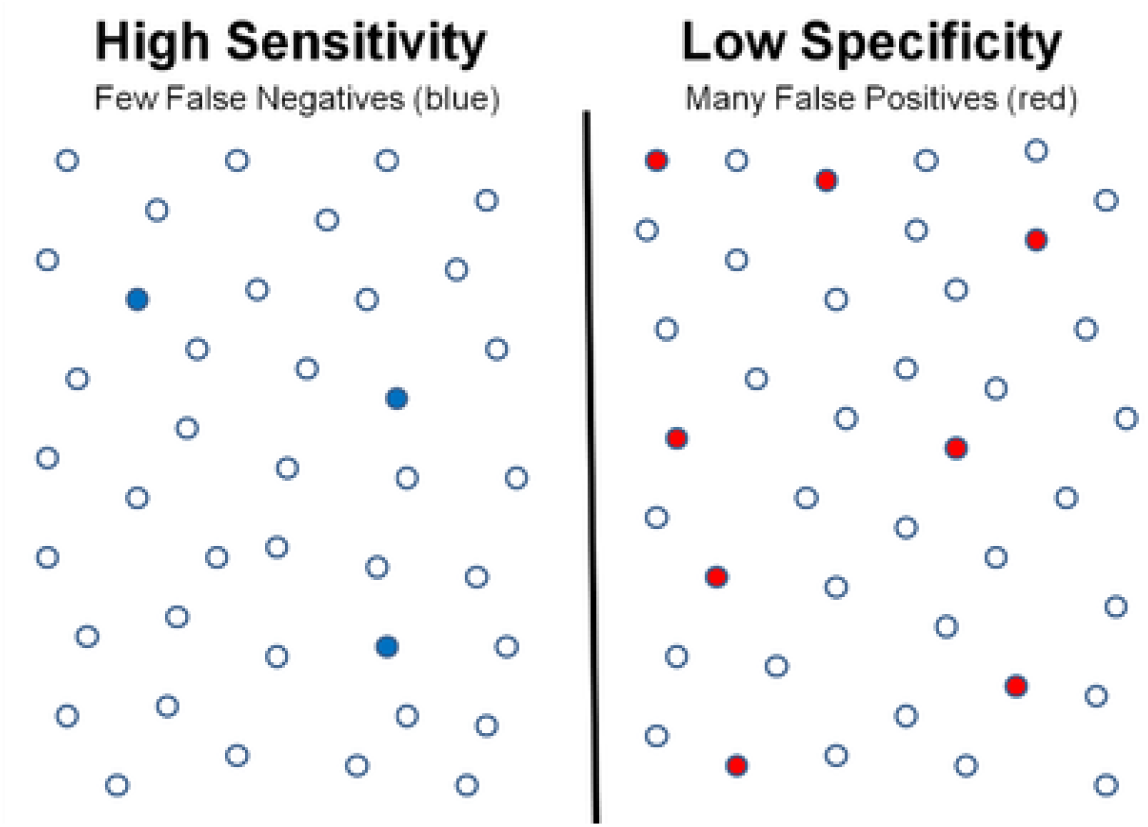

The figure above is a visual representation of False Negatives and False Positives in a particular dataset. The few numbers of these false predictions signify a strong model.

#### 4.1.3 Accuracy

Outside of specificity and sensitivity, accuracy remains a metric that is the most common in deep learning model evaluation. It is simply calculated as the ratio of correctly identified cases (that is, #True Positives + #True Negatives) and the total number of cases.

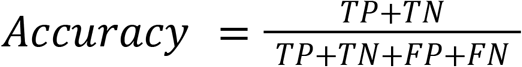

#### 4.1.4 ROC Curve

Although sensitivity and specificity are strong metrics for evaluating a deep learning model, they lack the visual aspect to make more conclusions. A ROC curve is created by plotting sensitivity against 1-specificity of various cases [8]. Typically, these graphs contain a linear line y=x as a reference to show how the model is different from a random classifier. A strong classifier aims to maximize the area under the ROC curve (AUROC). Evidently, a curve that hugs the y-axis and upper x-axis will have the largest area under the curve and a sign of a good model.

The labeled figure below is an example of a ROC curve and calculated AUROC value.

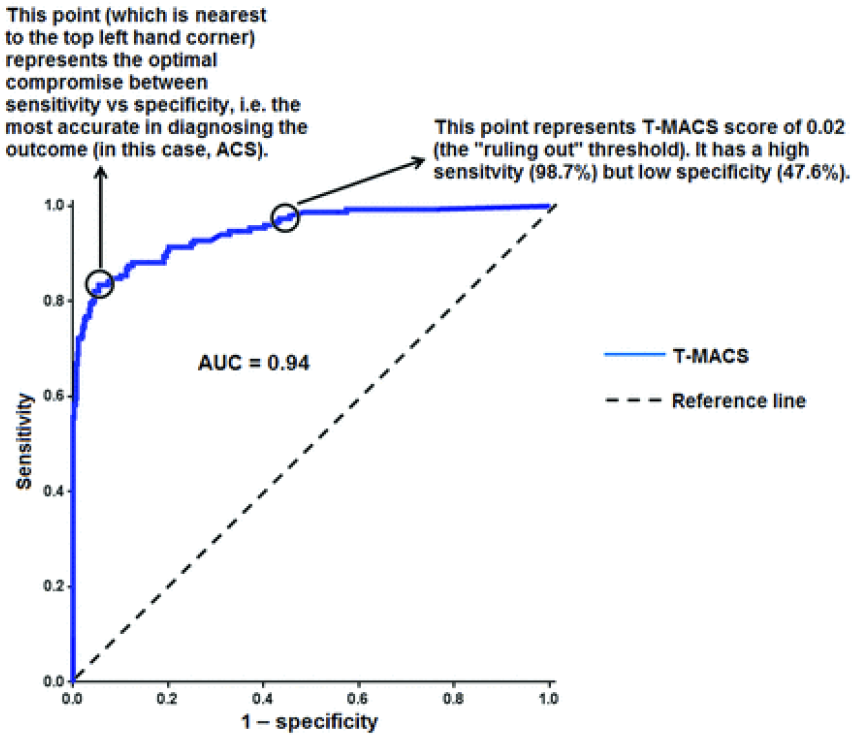

### 4.2 Summary

Using the hybrid model of VGG-16 and Inception v3, the model had an accuracy of 87.5% (remarkable accuracy for deep learning models in the field of medical imaging) and an AUROC of 0.929. Of course, like most deep learning models, there were problems that needed to be addressed during training to legitimize results. Learning rate drop-offs were added to prevent overfitting as well as image augmentation.

### 4.3 Statistics

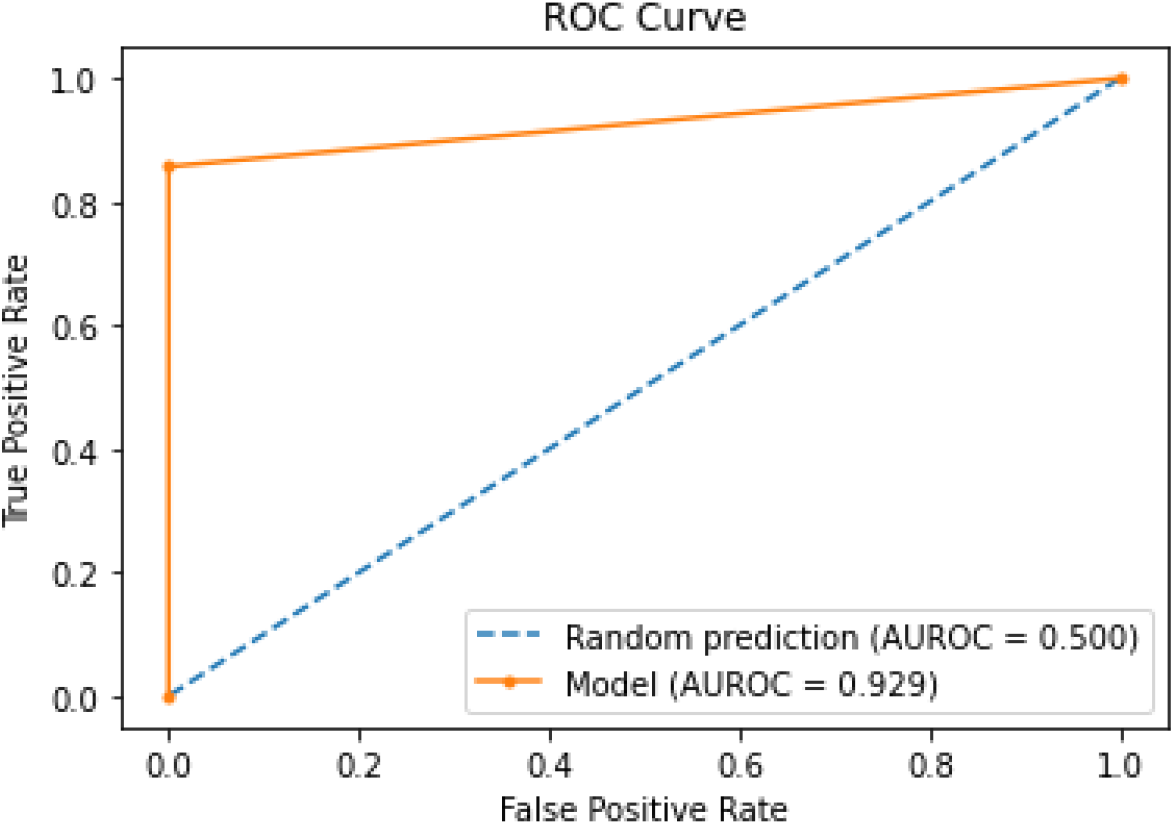

The ROC Curve above is graphed with a linear line of the equation y=x to demonstrate the accuracy difference of the hybrid model and a random classifier. Generally, a strong model (that isn’t overfitted) has an area under the ROC curve of 0.9 < AUROC < 0.95. The AUROC value shows that our model did not exploit the unbalanced data and work as a binary classifier but rather that it had hyper tuned parameters for accurate image analysis.

#### 4.3.2 Incorrectly Identified Case Analysis

Despite ROC statistics being relatively high in the deep learning context, it is still important to address the reasons for inaccuracy in this metric, the main one being the size of the dataset. The TDID dataset is very skewed in terms of positive and negative cases and even though this is accounted for using pre-generated weights, it’s no surprise that a model would be trained to favor the majority class. This problem can be avoided with a larger dataset that can be provided through a clinical dataset from a radiologist.

#### 4.3.1 Training

The hybrid model was trained for 10 epochs with a batch size of 8. Batch size is the number of image samples that will be passed through to the neural network. Backpropagation is run after every batch hence the larger the batch size, accuracy is compromised but training time increases greatly. Because of the small dataset size and the computing power of the PC this model is being trained on, we could afford to have a very small batch size and still receive results in a timely manner. As there is no select way to choose the perfect amount of epochs and batch size for any model currently, a lengthy process was taken to fine-tune these hyperparameters to produce the most reliable model.

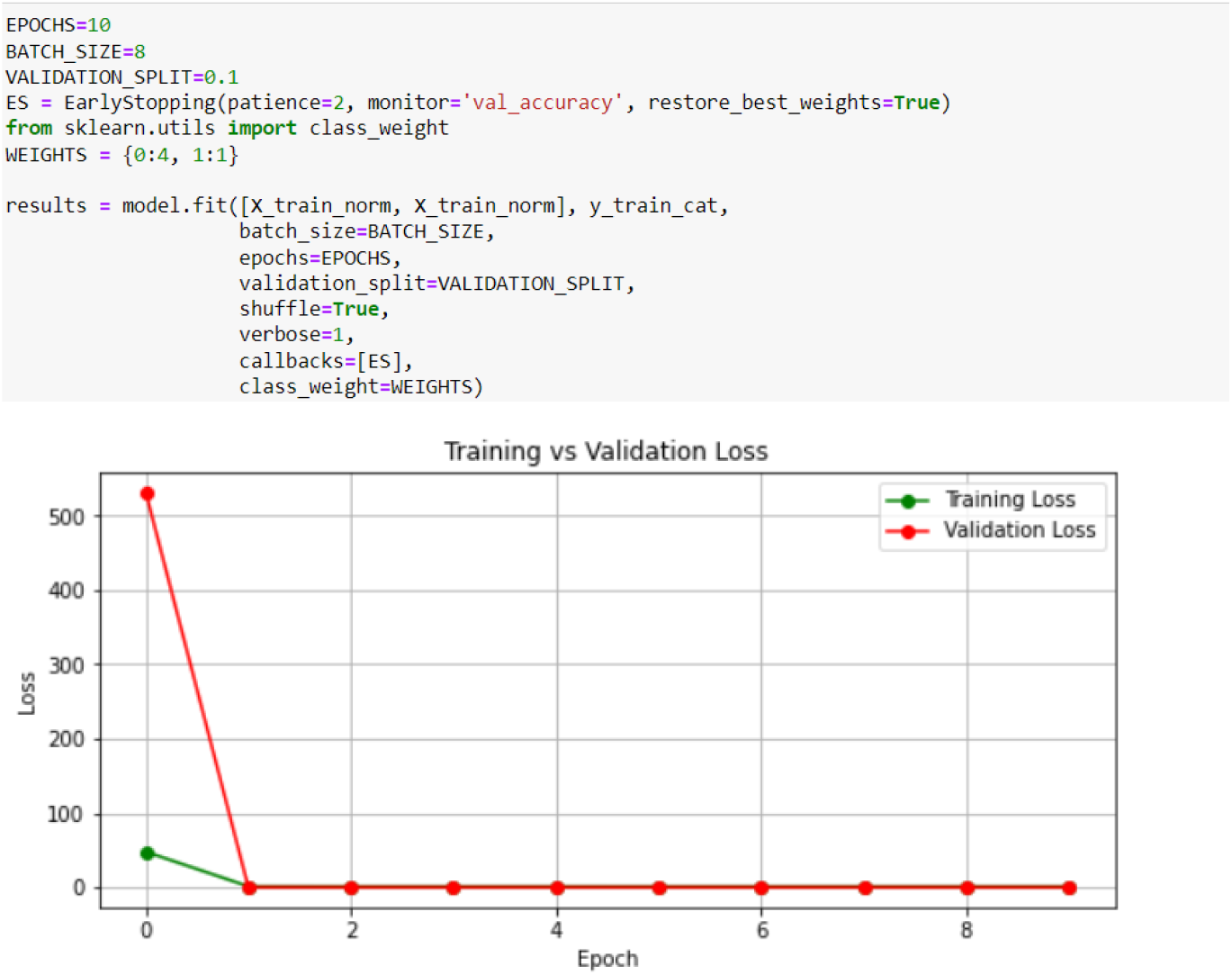

The graph above shows the training process of the model. Note: though the model appears to have zero loss after the first epoch, those points represent very small numbers that are hard to represent on the given axis.

## 5 DISCUSSION

The application of 2 pre-trained CNN models contributed to the strong results of the combined network. Because both of these models had been created for the basis of general image analysis, with Inception v3 being more tailored for medical imaging, this made the identification of thyroid nodules far more accurate. Additionally, the adoption of pre-generated weights to accommodate for data skew prevented the model from overfitting and simply favoring the majority class. Unbalanced data was accounted for by applying weights to certain nodes prior to training the model. The choice of binary cross-entropy was the most effective for this classification task between 2 classes. With a larger dataset, the model can become significantly more refined and reputable. The addition of normal images would also add more depth to the model rather than it being simply a binary classification. Overall, the model illustrates that a convolutional neural network can serve as an accurate determiner of malignant/benign thyroid nodules from an ultrasound in place of (or to further support) a radiologist. Hopefully, the model eventually becomes a viable resource for medical experts diagnosing thyroid cancers.

## Data Availability

All data produced are available online at
http://cimalab.unal.edu.co/?lang=en&mod=program&id=5

## 6. ACKNOWLEDGEMENTS

I would like to thank my parents for always showing their love and being there to support me. I am also very grateful for Dr. Aaron Kornblith’s mentorship in the paper-writing process. Finally, I would like to formally thank JSHS for hosting this competition and creating an outlet for young researchers to showcase their work for a good cause.

